# An Endpoint Adjudication Committee for the Assessment of Computed Tomography Scans in Fracture Healing

**DOI:** 10.1101/2023.06.04.23290949

**Authors:** Chloe Elliott, Ethan Patterson, Brenna Mattiello, Adina Tarcea, Bevan Frizzell, Richard E. Walker, Kevin A. Hildebrand, Neil J. White

## Abstract

The use of endpoint adjudication committees (EACs) has the potential to reduce subjectivity and potential bias in clinical research trials and contribute to a higher quality of research. In a recent randomized control trial (RCT), we used serial computed tomography (CT) imaging to visualize fracture healing of the scaphoid as a primary outcome. The scaphoid bone poses a challenge in the diagnosing of fractures and non-unions due to its complicated shape. An EAC was created to increase the quality of the data and the validity of our findings. While an adjudication process has long been proposed and described for X-rays, this study outlines a rational approach to CT scan adjudication for bone fracture healing. A total of 364 scans were acquired in the RCT and of these, 101 were adjudicated for a binary endpoint of union vs. non-union. The application of EACs such as described in this paper is a useful tool in orthopaedic research requiring the adjudication of fracture healing as a study outcome.

## Introduction

A randomized controlled trial is only as good as its primary outcome. In studies regarding bone healing, fracture union is often a desired trial outcome, albeit a historically unreliable one. Fracture healing determined by radiographs or CT scans lends itself to a large degree of subjectivity. Standard definitions of union, delayed union, non-union and partial union remain elusive. Dozens of studies have highlighted the poor intra- and inter-observer reliability of assessing radiographs for fracture union [1-8]. Vannabouathong et al. [1] observed this knowledge gap in the assessment of long bone fractures. They described and advocated for an endpoint adjudication committee (EAC) to be used when determining fracture union as a primary outcome in research. The interpretation of fracture union is subject to the individual opinions of professionals and must be cooperatively decided upon by a committee. Bhandari et al. [2] found that in the case of tibial shaft fractures, surgeons’ definitions of delayed union ranged from one to eight months. This disparity is crucial in determining the primary outcomes of clinical trials. An adjudication committee is highly recommended to determine subjective assessments, such as bone healing [3].

Adjudication is the process by which a debatable topic is deliberated by a panel of experts. The main goal of an EAC is to ultimately derive the best possible answer and remove uncertainty, therefore, significantly increasing primary outcome reliability. The current literature displays the significance of EACs and their value in studies involving cardiovascular, neurological, and fracture assessment [9], and in orthopaedics has been predominantly proposed for the assessment of long bone fracture healing [1]. EACs are recommended by the Food and Drug Administration (FDA) to be implemented in any form of clinical research that is at risk for subjectivity that may vary primary outcomes [5]. Dechartres et al. [3] investigated the use of adjudication committees in bone healing and asserted that they have a direct impact on the study result with up to a 20-30% change in union status after completing the adjudication process. An EAC tasked with determining union status of a series of fractures should be comprised of specialized surgeons and subspecialty trained musculoskeletal (MSK) radiologists. The use of adjudication in cases where primary outcome measures are subjective, as the FDA suggests, would benefit in terms of accuracy and ability for reproducible results by other parties.

The scaphoid is the most commonly fractured carpal bone and has a high rate of non-union [10]. Scaphoid fractures account for 60% of all carpal fractures and 2-3% of all fractures [11]. Due to the complex geometric shape of the scaphoid, judging union based on radiographic imaging is challenging and involves a high rate of subjectivity [12]. The diagnosis of scaphoid non-union (SNU) is usually given months or years after the initial injury and is commonly identified through standard radiographic assessment including wrist and scaphoid views [12]. X-ray examination, however, allows for accurate diagnosis of scaphoid fractures in only 70-80% of cases [11]. CT imaging is used to help diagnose scaphoid fractures when presented with positive clinical symptoms and negative x-rays. Considering this, Singh et al. [7] conducted a study that suggests a prominent level of difficulty in deciphering partial union of scaphoid fractures without the use of CT scans. They further highlighted that the difficulty in the assessment of union may lead to poor injury management. Matzon et al. [13] complemented this finding by describing the increased difficulty in CT evaluation after internal fixation. The challenge to adequately diagnose scaphoid union and non-union can cause long-term disability and inhibit the daily activities of patients. A more accurate diagnostic approach to scaphoid fractures would allow for these complications to be identified sooner, improve patient outcomes, and add credibility within research.

The senior author of this manuscript (NJW) conducted a Level I double-blinded randomized controlled trial assessing surgical SNUs and the effects of LIPUS as an adjunct to surgery (the SNAPU Trial). The primary outcome of the study was time to union of the scaphoid following surgery for an established non-union and the post-operative application of a LIPUS unit.

Patients were randomized to either ‘active’ or ‘sham’ units. Union status was determined with the use of serial CT scans. The initial trial design prescribed a fellowship-trained hand surgeon and MSK radiologist to independently read each CT scan and assess union using quartiles of percent union and a cutoff of 50% bridging for the definition of union (see S1 Appendix for study overview). It was soon realized the degree of variability between surgeons’ and radiologists’ interpretations necessitated a more standardized protocol to maintain the quality of the trial and ensure the best possible primary outcome. After conducting a literature review to find a pre-existing standardized tool for assessing union status on CT scan, the investigators identified a gap in the literature and formed their own standardized adjudication process. The purpose of this paper is to describe this adjudication process for reference, critique, and use by other researchers.

## Methods

The SNAPU trial was registered at clinicaltrials.gov (NCT02383160) and local and subsite ethics were obtained (REB13-0849). Recruitment for this trial took place from 2014 Jul to 2020 Feb and the last date of follow-up was 2022 May 19. The primary outcome of the SNAPU trial was time to union of the scaphoid after surgical fixation for established non-unions. We hypothesized that subjects randomized to a functional LIPUS device have a shorter time to union than subjects randomized to a placebo LIPUS device. The patients were instructed to use the LIPUS device once daily for 20 minutes until either union or non-union was declared. Serial CT scans were collected at 8 +/-2 weeks, 12 +/-2 weeks, and then every 4-6 weeks thereafter until either union was reached or a recurrent non-union was declared. All subjects were asked to continue to attend follow-up appointments until two years after the index procedure.

Over the course of the trial, 364 CT scans from 142 participants were accumulated (average 2.56/patient). These scans consisted of coronal and lateral two-dimensional serial views. Exact CT scan acquisition and reconstruction parameters used can be found in S2 Appendix. For each CT scan, both the treating fellowship-trained hand surgeon and MSK radiologist reported union status based on the following six classifications: 0%, 1-24%, 25-49%, 50-74%, 75-99% and 100%. The scaphoid was considered united once a threshold of 50% union was met as outlined by Singh et al. [7]. This double assessment process was designed to ensure an accurate assessment of union. However, there were three outcomes that came from this dual interpretation:

1. The treating surgeon and MSK radiologist agreed on the union percentage and no further action was required.
2. The treating surgeon and MSK radiologist disagreed on the percent quartile but differed by only one quartile (I.e., 0-24% and 25-49%) and it did not cross the critical threshold of 50% union. In this case, the CT scan was not adjudicated. The authors termed this a “minor discrepancy.”
3. The treating surgeon and MSK radiologist disagreed on the union status. This was declared as a “major discrepancy” and the CT scans in question were subsequently flagged for adjudication. A major discrepancy was defined as any time the surgeon and radiologist disagreed by more than one quartile (i.e., 25-49% vs. 75-99%) or more than one quartile away from the extremes 0% and 100% (i.e., 0% vs. 25-49%) or a single quartile that crossed the critical threshold of 50% union. Note that in the SNAPU trail, the classifications also included 0% and 100% which was not used in the adjudication process.

### Adjudication Committee Members and Charter

To address these discrepancies between professionals, the investigators implemented an EAC, using previously established guidelines regarding adjudication of radiographic imaging [2, 3]. The investigators designed an efficient adjudication protocol that could be effectively adapted to CT images in the SNAPU trial.

**Step 1: Identifying committee members and roles:**

- The EAC consisted of fellowship-trained hand surgeons and MSK radiologists
- There were three elected members of the adjudication committee to allow for a consensus to be reached on each CT scan
- Eligible individuals had to meet the following criteria: a minimum of 10 years of clinical experience and fellowship-trained in either hand and wrist surgery or musculoskeletal radiology
- The panel consisted of both MSK radiologists and surgeons, indifferent and unbiased toward study outcomes, with the single goal of assessing CT scans with major discrepancies to come to an agreement on the union status of the scaphoid at a given time point
- An experienced MSK radiologist was selected to be the chair of this EAC

The project manager identified all discrepant CT scans and created a de-identified worksheet (IMPAX for local images, Aycan/ResMD for sub-sites). The project manager also created spreadsheets that recorded outcomes and had available spaces for notes. The primary investigator (NJW) observed, organized, and facilitated the adjudication meetings, but had no role in CT image reading or decision-making during the adjudication process. The project manager (BM) and the principal investigator of the SNAPU trial (NJW) had access to identifying patient information during the adjudication process, but panel members did not. Other research team members involved in the SNAPU trial (CE, EP, AT) also had access to information that could identify individual participants.

**Step 2: Charter of Adjudication (See S3 Appendix for more details)**

The EAC was created by the selected chair. The charter created for the SNAPU trial EAC outlines the process, and rules, of the adjudication process. The rules for the committee included the following:

- Reviewers had access to both X-ray and CT images from each time point on every subject;
- All serial imaging leading up to the point in question may be reviewed
- If a single participant had more than one CT scan requiring adjudication they were evaluated separately. The first time was adjudicated with earlier imaging available only and later (not sequentially) the second time point was adjudicated as a discreet event.
- Time from surgery to adjudicated CT scan was known to the committee
- The committee was blinded to the original reading on the CT scan as well as functional vs. sham device status
- Reviewers did not read the original radiology report for any interval image
- The hardware (screw or k-wires) is NOT included in the calculation of cross-sectional area
- The goal of the committee was agreement on a specific quartile between all three reviewers

**Step 3: Adjudication Meetings**

Before the start of the adjudication panel, the process was outlined by the committee chair to ensure understanding of the following:

- Confirmation that all members accept methodology and the grading for determining percentage union by CT scan (Table 1).

**Table 1.** Quartiles of Union Status Identification
- Project manager would provide spreadsheets with discrepant cases to review
- The reviewer would access cases through IMPAX and AYCAN using existing credentials.
- The panel members were sent the charter for review, feedback and approval, and the panel members would confirm no conflicts of interest to the study coordinator.

| Percent Union (%) | 0-24 | 25-49 | 50-74 | 75-100 |
| --- | --- | --- | --- | --- |
| Classification | Non-Union | Non-Union | Union | Union |

The group held two separate formal adjudication meetings (2020 Oct 26 and 2021 Jun 10), totaling 4-5 hours for each meeting. A unique aspect of this process is that it was performed during the pandemic and it incorporated the use of remote access to imaging systems in combination with digital video communication platforms. The first step of the process was to identify the flagged CT scan as a major discrepancy and identify the number of weeks from surgery. The EAC members were encouraged to take notes while reviewing the images. Each scan was viewed independently by the EAC members for 2-3 minutes. The group then was brought together to discuss the images for another 5-7 minutes. The group could refer to imaging at any other time point before the flagged images on request. The committee was limited to a total of 10 minutes per CT scan and took an average of 7 minutes to arrive at an agreement. Committee members confirmed to the project manager once a final decision was agreed upon, and both the principal investigator and the project manager recorded the final decision. Where there was unanimous agreement, these data points were entered as final, but annotated as adjudicated. For the cases where a major discrepancy remained, the committee was set to meet further and review to reach a consensus. The SNAPU trial used the final decision by the board of adjudicators for all statistical analysis.

## Results

In total, 364 CT images were assessed by the treating surgeon and an MSK radiologist. Before adjudication, the surgeon and radiologist agreed on the exact quartiles (Table 1) on 153 of the CT images and found minor discrepancies in 114 scans. The EAC assessed 101 CT scans which were collected across 5 different study sites. Seventy-five (74.3%) of these CT scans were defined as major discrepancies (Table 2) and the remaining 26 CT scans were adjudicated due to missing data or as a random selection to validate the other groups (S4 Appendix). Of these 75 major discrepant cases, the final reading of the CT images by the EAC matched the initial reading of the MSK radiologist in 42 cases (56.0%) and the treating hand surgeon in 25 cases (33.3%). The remaining 8 CT images that were adjudicated to agree with both (6) or neither (2) the surgeon and/or MSK radiologist (10.7%). The EAC resulted in a final diagnosis of union in 41 (40.6%) cases and in non-union in 60 cases (59.4%) (Fig 1).

**Table 2.** CT Scans with Minor, Major and No Discrepancies.

| Discrepancy Grade | Minor | Major | None | Incomplete data | Total |
| --- | --- | --- | --- | --- | --- |
| Number of CT scans | 114 | 75 | 153 | 22 | 364 |
| Number of CT scans Adjudicated | 0 | 75 | 4 | 22 | 101 |

**Fig 1.**
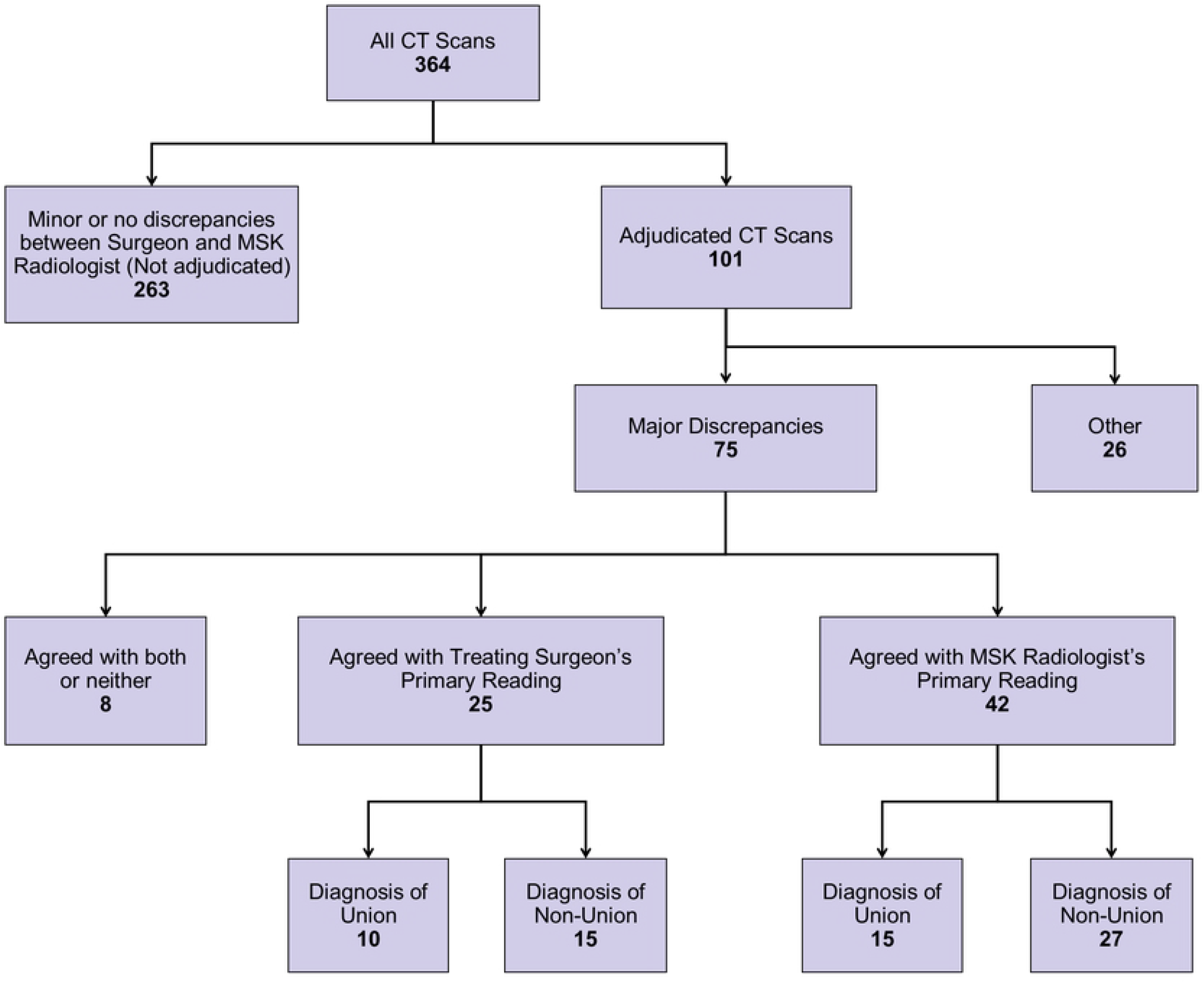
SNAPU imaging CONSORT Diagram.

## Discussion

### Success of the EAC

CT scans are considered the gold standard for measuring union in scaphoid fractures for both research and clinical settings [1]. However, as highlighted by the SNAPU trial and other research, even with CT imaging, trained and experienced clinicians do not always agree on a diagnosis [13].

The primary outcome of the SNAPU trial was time to union as defined by 50% osseous bridging. The EAC changed the final union classification of 83 CT scans. The use of an adjudication committee for nearly one third of the total CT scans reviewed in the SNAPU trial underscored the vital role that the EAC played in determining the trial’s results. The addition of an EAC in the SNAPU trial allowed for more accurate union assessment, and therefore provided a more reliable and consistent evaluation of scaphoid union. This can lead to fewer errors and more precise diagnoses and improving credibility of research. The process of adjudication in the radiological assessment of CT images can be implemented in clinical studies and have a positive impact on data analysis of primary outcomes.

The process of adjudication based on quartiles and both “minor” and “major” discriminators obviated the need to adjudicate all CT scans creating an efficient and logical process to arrive at the most accurate determination of union status. All CT scans in the SNAPU trial had a union status that was either agreed upon by an MSK radiologist and hand surgeon during the first pass or adjudicated and agreed upon by the panel on the second pass. To our knowledge, a process for adjudication of CT imaging has not been described in orthopaedic literature. While our process has limitations, it satisfies a balance between accuracy and feasibility.

Much of the orthopaedic literature is dependent on radiographic assessment and as we delve deeper into the 21^st^ century, researchers are depending more on advanced imaging to make accurate assessments. Regardless of the imaging modality, results and diagnosis are reliant on interpretation from an expert. This opens the door to error, due to limitations in accuracy and removal of bias. We propose that EAC’s serve as the gold-standard in high quality research when the primary outcome has any degree of subjectivity.

### Limitations

We recognize as pioneers of adjudication committees for CT images, that the design presented here is not without faults. One limitation is that review of the CT scans was limited to a maximum of 10 minutes each to provide efficacy in this process. This could possibly influence the decisions made by the clinicians as the diagnosis may have been different if they had been given more time to read and analyze the images. Images were adjudicated for an average of 7 minutes, but more time, such as a 15-minute limit, for example, would ensure that the reading would not be rushed. It was necessary to balance accuracy with feasibility and efficiency.

Another weakness is that not every CT scan obtained from the trial was adjudicated. Only major discrepancies were reassessed (union vs. non-union or two or more grades difference) by adjudication. This was for reasons of efficiency as we deemed it was not practical to ask professionals to adjudicate every image. The multiple study sites in the SNAPU trial created a third limitation. There were multiple CT scanners and CT scanning protocols used allowing for varying imaging quality and reformatting methods. This limitation was present because not every site was able to use the exact same CT parameters for all imaging. However, this is reflective of practice in the real world and of the fact that surgeons and radiologists are required to interpret varying CT images in their day to day practice. To reduce discrepancies caused by this challenge, using the same CT scanner parameters as well as radiologist personnel would help with consistency in image acquisition.

## Conclusion

The analysis in this paper confirms that the adjudication process had an impact on the assessment of scaphoid fracture union on CT scan during the SNAPU trial. The process allowed for imaging to be reviewed subjectively by a team of experts to define union in cases where discrepancies were present, allowing for a more valid analysis of the LIPUS device in scaphoid fracture healing. The adjudication process in the analysis of serial CT scans in fracture healing trials is a novel and pivotal step to the assessment, treatment, and outcome of fracture healing. EACs should be used routinely in orthopaedic trials where the endpoint is radiographic union.

## Data Availability

All relevant data are within the manuscript and its Supporting Information files.

---

**S1 Appendix. Introduction to SNAPU Trial**

**S2 Appendix. CT Scan Protocol**

**S3 Appendix. The Endpoint Adjudication Committee (EAC) Charter**

**S4 Appendix. The “Other” Cases Adjudicated by EAC**

